# Diagnostic accuracy of B scan ultrasonography for posterior segment eye disorders in a tertiary care setting in Eastern India

**DOI:** 10.1101/2020.08.13.20173898

**Authors:** Maheswar Chaudhury, Bikash Parida, Sandeep Kumar Panigrahi

**Affiliations:** Department of Radiology, IMS and SUM Hospital, Siksha ‘O’ Anusandhan deemed to be University; Community Medicine department, IMS and SUM Hospital, Siksha ‘O’ Anusandhan deemed to be University, Bhubaneswar

**Author notes:** Corresponding author: Dr. Bikash Parida, Assistant Professor, Department of Radiology, IMS and SUM Hospital, Siksha ‘O’ Anusandhan deemed to be University, Bhubaneswar, Odisha – 751003.

**Keywords:** ultrasonography, posterior eye segment, sensitivity and specificity, predictive value of tests

## Abstract

**Introduction:** Ophthalmic ultrasound (USG) produces real time high resolution images of the eye and orbit. It can categorize and predict the location of pathology in the posterior chamber of eye very well. It is useful even in pre-operative evaluation and diagnosing posterior segment eye disorders. However, the diagnostic accuracy has usually not being studied thoroughly, with special emphasis to its probability of predicting posterior-segment eye disorders using B-scan USG.

**Objectives:** To find out the prevalence and pattern of posterior segment disorders using B-scan ultrasonography, and to find its diagnostic accuracy.

**Materials and Methods:** The study was prospective in nature and conducted in the department of radiodiagnosis and ophthalmology of a tertiary care center of Eastern India. Patients referred to the radiology department for ruling out intra-ocular pathology using B-scan ophthalmic ultrasound were included in the study, irrespective of any age and gender. Data were captured on an excel sheet and analyzed using Stata 12.1 SE.

**Results:** The mean age of 84 study participants was 37.4 ± 19.5 years, with maximum in between 40-50 years. Males were more (72.6%). 50% presented with low vision, and most commonly associated with cataract (45%). Prevalence of posterior segment eye disorders was 13.1%. Sensitivity and negative predictive values were 100%. Post-test probability was 95.5%. Accuracy was however very less (39.3%).

**Conclusion:** Using B-scan ultrasonography for pre-operative assessment and confirmation of diagnosis increases the probability of detecting presence or absence of posterior segment pathology. Absence of posterior segment disorder using this is also very helpful in ruling out disease entirely. It also a very high sensitivity and hence can be used even in rural health centers.

## Introduction

Eye is situated in the anterior part of the orbit embedded in fat with the Tenon’s capsule separating it from the orbital wall. Posterior segment of the eye, which constitutes the major portion (5/6^th^ of the eye) is composed of vitreous cavity, retina, choroid, sclera, episclera, and occasionally subluxated lens.

Cystic composition and even superficial location makes the eye an ideal organ for ultrasonographic (USG) examination with unique acoustic advantages being offered by its ultrasound imaging. The anechoic vitreous acts as a natural contrast for ultrasound purposes. Ultrasound becomes the most practical method of obtaining images of the posterior segment when light conducting media are opaque. It is most commonly employed prior to vitrectomy also. Ophthalmic USG may be of A-type or B-type. A-scan can be used for biometric calculation, while B-scan for quantifying the reflectivity of lesions in the eye and orbit. A 20 MHz probe is used for lens and vitreous while 50 MHz probe for anterior segment (also known as ultrasound bio-microscopy). Routine ocular scanning is done with B-scan using 7.5 MHz or above frequency transducers. ^1,2^

Ophthalmic USG offers multiple advantages over Computerized tomographic (CT) scan and Magnetic Resonance Imaging (MRI). It produces real time images of the eye and orbit with a very high resolution (0.1 to 0.01). Multiple cross-sectional and radial cuts can be rapidly obtained at the bedside or even in an operating room, as it is portable. It is less expensive and without radiation hazards. It can categorize and predict the location of pathology in the posterior chamber of eye very well.^3^

Indications of ophthalmic ultrasound include evaluation of a broad range of conditions like suspected intraocular tumor, localization of the foreign body in eye, eye trauma conditions, examining the vitreous, opacity in conducting media of eye making ophthalmoscopy difficult, etc. B-scan USG finds a good place in evaluating these cases, and has proven to be a rapid, reliable, safe and cheap investigation. It is useful even in pre-operative evaluation and diagnosing posterior segment eye disorders.^2^ However, the diagnostic accuracy has usually not being studied thoroughly, with special emphasis to its probability of predicting posterior-segment eye disorders post USG. Thus in this study the authors wanted to find out the prevalence of posterior segment eye disorders and diagnostic accuracy of B-scan USG for posterior segment eye disorders at a tertiary care center in cases referred to radiology department.

## Objectives

1. To find out the prevalence and pattern of posterior segment disorders among patients referred to radiology department using B-scan ultrasonography.
2. To find out the diagnostic accuracy of B-scan ultrasound in the diagnosis of posterior segment ophthalmic disorders among patients referred to radiology department.

## Materials and Methods

The study was hospital based and prospective in nature. It was conducted at a tertiary care center of Eastern India from 2007 to 2009 jointly in the department of radiodiagnosis and department of ophthalmology. Patients who presented to the ophthalmology department with suspected intra-ocular pathology, or in conditions where fundoscopy could not be carried out such as blunt trauma to the eye, opaque ocular media, etc., were referred to the radiology department for B-scan ophthalmic ultrasound. Their consent was taken and they were included in the study.

Inclusion criteria include patients with suspected posterior segment eye disorders (blunt trauma leading to opacity in media, intra-ocular pathology like retinal detachment, hemorrhage, tumour, retinopathy, etc.) irrespective of any age and gender. Exclusion criteria included patients who did not consent or agree for follow up, or who had been operated on the eye for similar conditions in the past, or had been already diagnosed using alternate methods such as fundoscopy, ophthalmic coherence tomography (OCT), magnetic resonance imaging, fluoroscopy, etc., which could possibly bias the selection process and also the interpretation.

Institute ethical clearance was taken before conducting the study, and patient confidentiality was maintained throughout the study. Written informed consent was taken before conducting USG. Data were entered in an excel sheet (Microsoft Excel 2010). They were cleaned for any missing values, and imported to Stata 12.1 SE for analysis. Data analysis were done for all the complete case records available. Qualitative data were presented as numbers with percentages, while quantitative data were represented with mean and standard deviation. Data were tabulated for calculating accuracy of B-mode USG. Diagnosis by conventional methods such as direct ophthalmoscopy was taken as gold standard for detecting posterior segment pathology. Cases where direct ophthalmoscopy could not be done such as presence of dense cataract, opaque media, etc. were grouped as cases where diagnosis were not done using gold standard methods. These were finally included under disease negative using gold standard method. Cases detected using both gold standard and B-mode USG were regarded as true positives (TP), detected by USG but not by gold standard as false negatives (FN), not detected by USG but detected by gold standard as false positives (FP) and not detected by both as true negatives (TN).

Accuracy of USG was calculated in comparison with gold standard method in terms of sensitivity (Sn), specificity (Sp), positive predictive value (PPV), negative predictive value (NPV) and accuracy. Likelihood ratios (LR^+^ and LR^-^) were calculated from sensitivity and specificity. Prevalence of the disease (posterior segment disorders) was calculated as proportion of cases diagnosed using gold standard (TP+FP) out of the total cases. Pre-test odds was calculated from the derived prevalence. Post-test odds was derived by multiplying pre-test odds with LR, while its probability was finally derived from post-test odds and Fagan’s nomogram.^4,5^

## Results

A total of 84 cases were referred to the radiology department from the ophthalmology for confirmation of posterior segment pathology or other conditions which could not be diagnosed using conventional methods (direct ophthalmology).

The mean age of the study participants was 37.37 ± 19.49 years (minimum 1 year, maximum 90 years). Maximum number of study participants were between 40-50 years at presentation (30 cases, 35.7%) (Fig 1).

**Fig 1:**
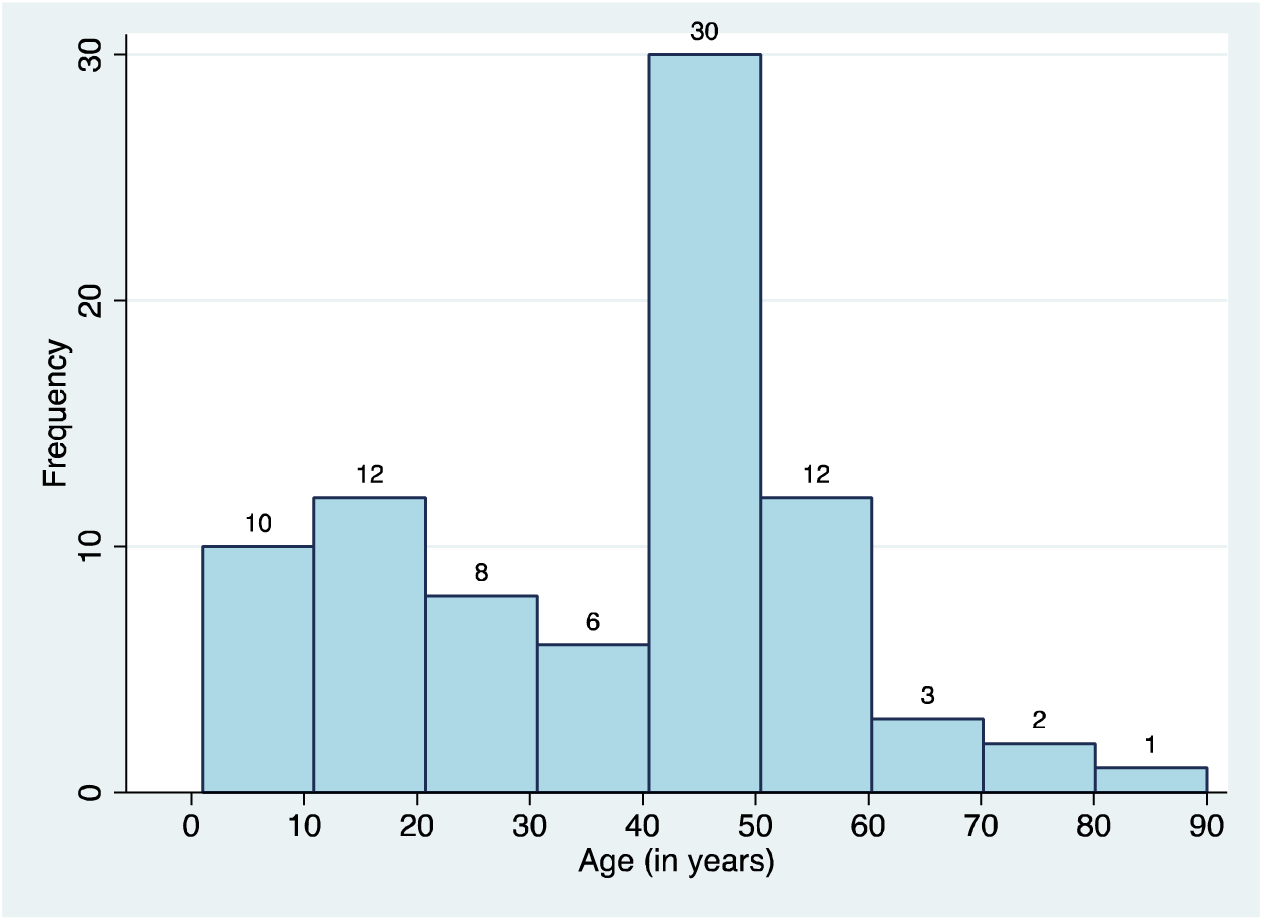
Age distribution of study participants (n=84)

The number of males among the study participants were 61 (72.6%) (Fig 2).

**Fig 2:**
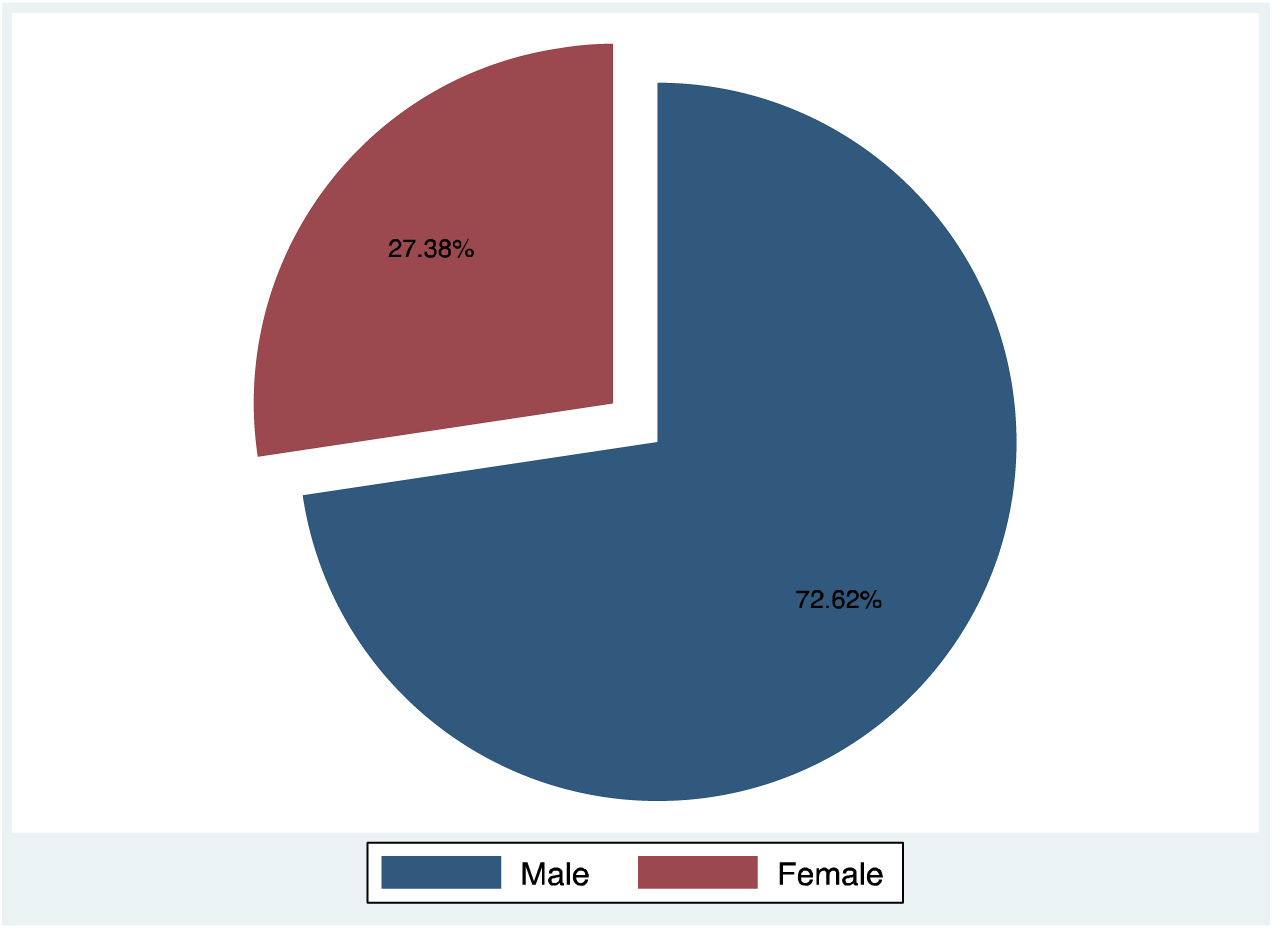
Sex distribution of study participants (n=84)

The common presenting symptoms among the referred cases were low vision (42 cases, 50.0%) followed by black spots in the visual field (9 cases, 10.7%) and pain around eyes (8 cases, 9.5%).

The most common condition that was found among the study participants was cataract/ corneal opacity (45 cases, 54%). Risk factors included raised intra-ocular pressure (8 cases, 10%). Diabetes and hypertension were however also the presenting general risk factors among the study participants in few cases (8% and 5% respectively) (Table 2). Of the 45 cataract cases, three had complete loss of vision, while rest had low vision (Table 1).

**Table 1:** Presenting symptoms among study participants (n=84)

| Presenting symptoms | Numbers | % |
| --- | --- | --- |
| Low vision | 42 | 50.00 |
| Black spots in visual field | 9 | 10.71 |
| Pain around eyes | 8 | 9.52 |
| Flashes of light | 7 | 8.33 |
| Blurring of vision | 5 | 5.95 |
| Leukocoria | 5 | 5.95 |
| Loss of vision | 3 | 3.57 |
| Bleeding from eyes | 2 | 2.38 |
| Shrunken eyes | 1 | 1.19 |

**Table 2:** Risk factors of study participants (n=84)

| **Risk factors** | **Numbers** | **%** |
| --- | --- | --- |
| History of trauma | 33 | 39.29 |
| History of suspected intra-ocular pathology | 3 | 3.57 |
| Presence of general risk factors | 11 | 13.06 |
| *Diabetes Mellitus* | 7 | 8.30 |
| *Hypertension* | 4 | 4.76 |
| Presence of ocular risk factor | 17 | 20.23 |
| *Increased intra-ocular pressure* | 8 | 9.52 |
| *Posterior synechia* | 6 | 7.14 |
| *Subluxated lens* | 2 | 2.38 |
| *Keratic precipitates* | 1 | 1.19 |

Posterior segment eye disorders could be ruled out to a good extent in cataract cases, trauma cases, suspected intra-ocular pathology and presence of leukocoria. It was best for confirming diagnosis of a posterior segment disorder in the presence of leukocoria (5 out of 5 cases, 100%) and cataract (7 cases out of 7, 100%), followed by suspected intra-ocular pathology (2 out of 3 cases, 66.7%). Of all the cases of cataract, posterior segment pathology was suspected in only seven cases.

**Table 3:** Cross-tabulation of posterior segment disorder diagnosis by gold standard methods and ultrasonography (n=84)

| USG Finding | Diagnosis |  |  |  |  |  |  |
| --- | --- | --- | --- | --- | --- | --- | --- |
|  | CD+VH | N | ND | RD | RM | TL | VH |
| **CD+VH** | 1 | 0 | 0 | 0 | 0 | 0 | 0 |
| **N** | 0 | 23 | 37 | 0 | 0 | 0 | 0 |
| **RB** | 0 | 0 | 0 | 0 | 5 | 0 | 0 |
| **RD** | 0 | 0 | 6 | 1 | 0 | 0 | 0 |
| **SL** | 0 | 0 | 0 | 0 | 0 | 1 | 0 |
| **VH** | 0 | 0 | 7 | 0 | 0 | 0 | 3 |

CD+VH-Choroidal detachment with vitreous hemorrhage; N-Normal; RB-Retinoblastoma; RD-Retinal detachment; SL-Subluxated lens; VH-Vitreous hemorrhage; ND-Not done; RM-Retinal mass; TL-Tilted lens

Prevalence of posterior segment disorder (using gold standard method) was found to be 13.1%. Sensitivity of B-scan USG was found to be 100%, as was also the negative predictive value (Table 5). Positive likelihood ratio was found to be 143, which increased the post-test probability to 95.5% (also confirmed using Fagan’s nomogram, fig 3)

**Table 4:**
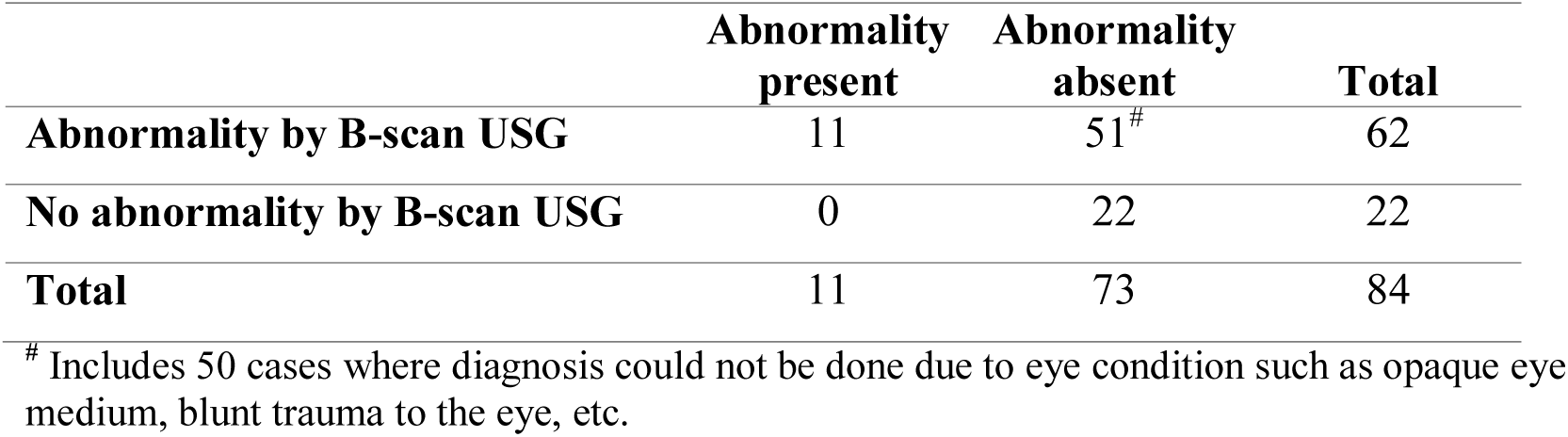
Accuracy of diagnosis using B-scan USG for posterior segment disorders (n=84)

|  | Abnormality present | Abnormality absent | Total |
| --- | --- | --- | --- |
| **Abnormality by B-scan USG** | 11 | 51<sup>#</sup> | 62 |
| **No abnormality by B-scan USG** | 0 | 22 | 22 |
| **Total** | 11 | 73 | 84 |

<sup>#</sup> Includes 50 cases where diagnosis could not be done due to eye condition such as opaque eye medium, blunt trauma to the eye, etc.

**Table 5:**
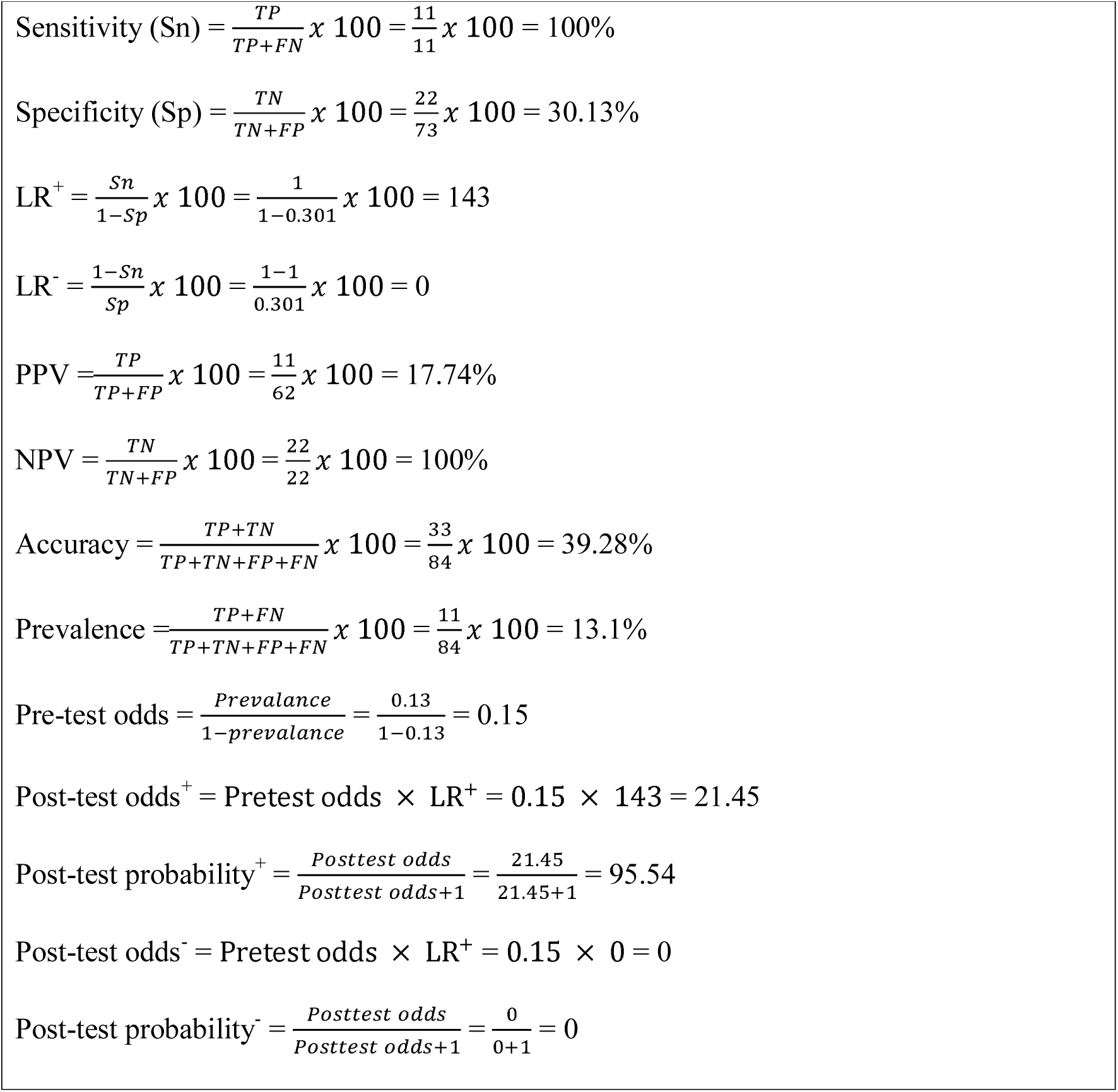
Calculations involved in diagnostic accuracy of B-scan USG among study participants (n=84)

**Fig 3:**
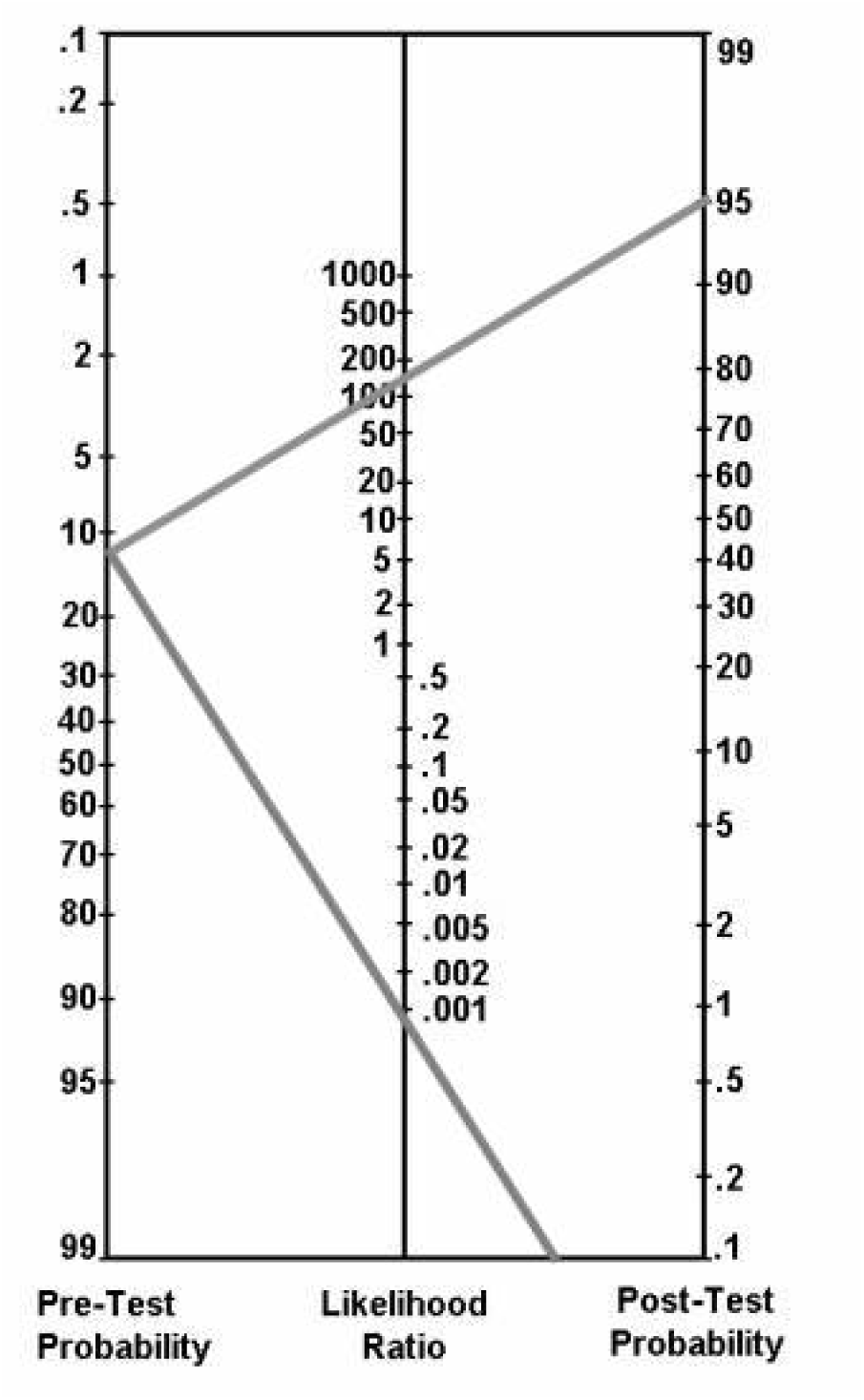
Fagan’s nomogram to detect post-test probability (LR^+^=143, LR^-^ = 0)

## Discussion

Ophthalmic ultrasound uses high frequency sound waves (>20 KHz) transmitted and received through a probe, and then converted into electric signals which are read on a monitor. The higher the frequency of ultrasound, the shorter is the wavelength. There is a direct relationship between wavelength and depth of the tissue penetrance (the shorter the wavelength, the more shallow is the penetrance). A shorter wavelength also gives a high resolution image. Ophthalmic ultrasound use around 10 MHz and thus produce detailed resolution images of the posterior segment of the eye. Sound travels faster in solid than in liquid mediums and eye is composed of both, an important aspect to be considered while dealing with the eye. Sound travelling from one medium to the other gets reflected back at the interface on to the probe, and this reflection is directly proportional to the density difference between the media. In B-scan USG, these echoes are represented as multiple dots forming an image on the screen. The stronger the echo, the brighter is the dot.^1^

Keeping the probe perpendicular to the area of interest gives a greater reflection on to the probe, and an inclined angle loses this intensity and clarity. Denser medium also end up with greater absorption, and thus compromising the resolution of the image. Presence of a dense cataract thus hampers the image resolution as compared to a normal crystalline lens, as also is a calcification mass. This may lead to no signal posterior to the medium, which is termed as shadowing. Pulse-echo system enables to compensate for this by amplifying the display through adjustment of the gain.^1^

Andreoli et.al., retrospectively reviewed ten years’ data (2000-2010) of 965 open globe injuries in Massachusetts where B-scan ultrasonography was used and found that the predictive value of B-scan USG was 100% for diagnosing retinal detachment and IOFB. Other diagnoses such as retinal detachment, disorganized posterior contents, haemorrhagic choroidal detachment, etc. were also found to have worse visual acuity at final follow-up. They concluded that B-scan USG can offer both diagnostic and prognostic information. It was also found to be useful for both surgical and medical management.^6^

In a study by Qureshi et.al. for pre-surgical planning of cataract patients done among 750 cataract cases, they concluded that if B-scan ultrasound is used as a diagnostic tool, the hidden posterior segment lesions can be detected prior to surgery. This if done routinely in pre-operative cases of cataract could help in surgical planning. If 2D B-scan is not helpful, then a 3D ultrasound can be opted for.^7^ Even for determining the integrity of posterior capsule in posterior polar cataract, this can be very efficiently used as shown by Guo et.al.^8^

In a study done by Sandinha et.al. on 58 patients to assess the accuracy of B-scan USG for acute fundus obscuring vitreous haemorrhage (FOVH) they found that B-scan U/S scan was highly sensitive in identifying the pathology in acute FOVH.^9^

In another comparative study done by Yang et.al. on alkali burn eyes, 56 cases were evaluated using both ultrasound bio microscopy and immersion 20 MHz B-scan USG, and compared with intra-operative findings. Compared with the intraoperative findings, the diagnostic concordance rate of B-scan appearance of lens was 100% (56/56), which was significantly higher than when done by an UBM 57.14% (32/56) (p <0.01).^10^

In a study conducted by Mohamed et.al. on 100 diabetic patients presenting with diabetic retinopathy using B-scan USG, it was seen that vitreous hemorrhage (VH) 42(66.6%), was the most important cause of low vision in patients presenting with poorly or moderately regulated HbA1c. Other reasons like asteroid hyalosis (AH), partial or total retinal detachment (PRD/ TRD), posterior vitreous detachment (PVD), and choroidal detachment (CD) were also found. They concluded that the non-invasive procedure of ophthalmic B-mode ultrasound can be used with minimum discomfort for DR complications linked with the visual outcome of a diabetic case.^11^

In a comparative study of bio-microscopy, B-scan USG and optical coherence computerized tomography done by Kivoca et.al., the posterior vitreous cortex was examined in 30 eyes among 30 patients presenting with macular pucker or macular hole, a day prior to a scheduled vitrectomy. It was seen that B-scan USG was the most reliable investigator-dependent prognostic method along with BM to clinically detect if the posterior vitreous cortex is detached.^12^

When fundus examination is not possible, ultrasound is known to be a better method for investigation of possible retinal tears. In a study done by Streho et.al., it was further found that B-scan USG is able to detect superior retinal tears predominantly than in other quadrants (71% cases among the 101 eyes of 100 patients studied).^13^

Case studies published have also shown that ultrasound B scan is an indispensable tool for the diagnosis of Ocular Cysticercosis, locating masses/ growths/ tumours even in complete cataract cases, diagnosis of microvascular changes associated with optic disc drusen, etc.^14-16^

In a study among 102 Sudanese adult patients with cataract done by Gareeballah et.al., sonographic findings showed posterior vitreous detachment (PVD) in 19.2%, vitreous hemorrhage of 0.98%, and complicated PVD in 1.96%. They also concluded that abnormalities of posterior segment should be evaluated by USG before surgery, especially cataract surgery.^17^

A sensitivity of 100% as found in the study indicates that all cases of posterior segment pathologies would be detected using B-scan USG, and almost none would be left undetected, thus making this a useful screening tool even in rural health care set-up. This has also been recommended by many authors.^2^ The specificity however, was found to be as low as 30%, and thus may not be an appropriate method for confirmation of diagnosis.

Likelihood ratios (LRs) are one of the best ways to know the diagnostic accuracy.^18^ It is the likelihood of a given test result in patient with a disease compared to the likelihood in a patient without the disease. The LR is used to assess and select a diagnostic test or sequence of tests. As compared to sensitivity and specificity, they are more or less stable with changes in prevalence of the disorder, and they can be used to calculate post-test probability of a disease using the test.^19^ In our study we had a very high LR^+^ (LR^+^ = 143) and a very low LR^-^ (LR^-^ = 0), which qualitatively are said to be excellent for prediction of positive and negative test results.^4^ The prevalence of diagnosing a posterior segment pathology at the tertiary care centre was found to be 13.1%, which means using gold standard technique posterior segment pathology would be detected in 13 cases out of 100 presenting referral cases from Ophthalmology department. However, using B-scan USG the post-test probability increased to 95.45%, i.e. the detection of posterior segment pathology would increase to 95 or more out of 100 such cases referred from the ophthalmology department.

The study had a limitation of not using a calculated sample size for determining the diagnostic accuracy of B-scan USG for posterior segment eye disorders.

## Conclusion

Using B-scan ultrasonography for pre-operative assessment and confirmation of diagnosis increases the probability of detecting presence or absence of posterior segment pathology. It also a very high sensitivity and hence can be used even in rural health centers. Absence of posterior segment disorder using this is also very helpful in ruling out disease entirely.

## Data Availability

Data is available with the authors and can be provided on request.

